# Pandemic excess mortality in Spain, Sweden, and Switzerland during the COVID-19 pandemic in 2020 was at its highest since 1918

**DOI:** 10.1101/2021.08.12.21261825

**Authors:** Kaspar Staub, Radoslaw Panczak, Katarina L. Matthes, Joël Floris, Claudia Berlin, Christoph Junker, Rolf Weitkunat, Svenn-Erik Mamelund, Matthias Egger, Marcel Zwahlen, Julien Riou

## Abstract

Estimating excess mortality allows quantification of overall pandemic impact. For recent decades, mortality data are easily accessible for most industrialized countries, but only a few countries have continuous data available for longer periods. Since Spain, Sweden, and Switzerland were militarily neutral and not involved in combat during both world wars, these countries have monthly all-cause mortality statistics available for over 100 years with no interruptions. We show that during the COVID-19 pandemic in 2020, Spain, Sweden and Switzerland recorded the highest aggregated monthly excess mortality (17%, 9% and 14%) since the 1918 influenza pandemic (53%, 33% and 49%), when compared to respective expected values. For Sweden and Switzerland, the highest monthly spikes in 2020 almost reached those of January 1890. These findings emphasize the historical dimensions of the ongoing pandemic and support the notion of a pandemic disaster memory gap.

**One-Sentence Summary:** During the COVID-19 pandemic in 2020, Spain, Sweden, and Switzerland recorded their highest monthly excess and all-cause mortality levels since the 1918 influenza pandemic, emphasizing the historical dimension of the ongoing pandemic.

---

During the COVID-19 pandemic in 2020, Spain, Sweden, and Switzerland recorded their highest monthly excess all-cause mortality levels driven by an infectious disease since the 1918 influenza pandemic. Although detailed mortality data from the last decades are easily accessible for most countries, only a few countries have continuous data available over the previous 100 years. Even fewer countries were unaffected by territory shifts or major losses of life during world wars. Since Spain, Sweden, and Switzerland were militarily neutral and not involved in combat during both world wars, they are rare cases where monthly all-cause death statistics are available for more than 100 years with no interruptions. These findings emphasize the historical dimensions of the ongoing pandemic and support the notion of a pandemic disaster memory gap.

Excess mortality compares expected and observed values; it is applied temporally as well as regionally. The expected number of deaths, or deaths predicted from those observed in previous years, derives from counterfactual thinking (*1, 2*) that occurs after an event and counters facts. The “expected” ideally captures what would have happened based on extrapolating observations from prior years. As such, excess mortality is increasingly used to quantify the COVID-19 pandemic’s overall impact in 2020 (*3*), including periods of excess mortality *(4)* and periods of lower-than-expected mortality *(6)*. Demographers have used excess mortality since the 1830s to describe monthly and seasonal mortality fluctuations (*4, 5*). In the 1850s, William Farr used expected versus observed crude death rates to identify places and populations that might benefit from sanitary interventions in England (*6*).

Europe has experienced several influenza pandemics over the last 140 years: “Russian flu” (1890); “Spanish flu” (1918); “Asian flu” (1957); “Hong Kong flu” (1968); “Chinese-Russian flu” (1977); and “swine flu” (2009) (*7*). The 1918 influenza caused the most deaths; consequently, it is the best documented and researched pandemic (*8*). In Europe, approximately 2.6 million excess deaths occurred during the 1918 influenza pandemic (*9*). An early application of the concept of excess mortality during a pandemic occurred when Switzerland’s federal authorities compared monthly deaths in 1890 with adjacent years in the 1890 influenza pandemic (*10*).

Estimates of excess mortality can provide information about the burden of mortality related to pandemics, including deaths that are indirectly linked to pandemics (*11*). In fact, the WHO estimated that in 2020 the total global deaths attributed to the pandemic is at least 3 million, representing 1.2 million more deaths than officially reported (*12*). Other global estimates showed that excess mortality substantially exceeded official deaths reported from COVID-19 (*1, 12, 13*). These estimates reinforced the relevance of excess mortality when examining a pandemic’s full mortality burden. However, at present, there are no studies comparing pandemics between countries based on uniformly structured data sets analyzed with a common approach.

To assess historical dimensions of the ongoing pandemic, we estimated age-specific monthly all-cause excess deaths for Spain, Sweden, and Switzerland for 2020 and other pandemic periods since the end of the 19^th^ century in chronological order. Spain, Sweden, and Switzerland are particularly well suited for longer-term contextualization of excess mortality due to pandemics based on available continuous data from 100 or more years. First, because these countries were militarily neutral and not involved in combat during both world wars, they were not simultaneously impacted by war-related excess mortality. Furthermore, collecting vital events and censuses continued without data quality issues such as those expected from countries at war. Second, since the end of the 19th century, these countries were not affected by significant territory shifts. And third, all 3 countries were heavily affected by the pandemic in 2020 (*1, 14*).

For these 3 countries, we analyzed the continuous series of officially reported deaths (all-cause) by month from the earliest available year (Spain 1908; Sweden 1851; Switzerland 1877) through the end of 2020. To calculate the expected values for a given year with uncertainty (e.g., 1918), we used deaths and total population counts from 5 prior years (e.g., 1913 – 1917). We excluded years with high pandemic mortality (i.e., 1890, 1918, 1957) to estimate expected values for the years following these pandemic years. We jointly modelled overall death counts by month and age group-specific death counts by year. We used negative binomial and multinomial likelihoods for the 2 counts, respectively. We used annual population counts as offsets (*15, 16*). We accounted for temporal trends and seasonal mortality variability by incorporating a linear calendar time trend and seasonality patterns with 4 flexible cosine and sine functions in the model (*17*). We implemented models in a Bayesian framework, allowing for full uncertainty propagation (see the Supplementary Material). We performed all analyses using R software version 4.1.0 (*18*) with the Stan software version 2.21 (*19*) interfaced by the rstan package (*20*).

During the pandemic in 2020, Sweden and Switzerland recorded the highest aggregated monthly excess mortality (9% [95% Credibility interval (CrI): 5 to 12] and 14% [10 to 19]) since the 1918 influenza pandemic (33% [27 to 39] and 49% [44 to 55]) (Table 1). This was also true for Spain (17% [12 to 22] versus 53% [48 to 59]), excluding Spanish civil war years, 1936-1939. In these countries, the percentage of excess mortality was thus 3–4 times higher in 1918 than in 2020. In the case of Spain, the civil war’s mortality impact is also of importance, yet difficult to quantify (*21*). Moreover, the peaks of monthly excess mortality in all 3 countries in 2020 were greater than any monthly excess mortality since 1918, including all seasonal flu peaks and heatwaves (Fig. 1). For Sweden and Switzerland, the highest monthly spikes in 2020 almost reached levels from January 1890: the peak of the 1890 influenza pandemic (when authorities hardly intervened with non-pharmaceutical measures).

**Table 1:** Characteristics (per calendar year) for the strongest 4 pandemics of the last 140 years in Spain, Sweden, and Switzerland.

| Year | Country | Population | Expected | Observed | Excess (CrI) | % Excess (CrI) | % Excess Pop |
| --- | --- | --- | --- | --- | --- | --- | --- |
| 1890 | Spain | - | - | - | - | - | - |
| 1890 | Sweden | 4,784,981 | 73,042 | 81,824 | 8,782 (4,830 to 12,435) | 12 (7 to 17) | 0.18 |
| 1890 | Switzerland | 2,960,800 | 58,355 | 61,805 | 3,451 (711 to 6,123) | 6 (1 to 11) | 0.12 |
| 1918 | Spain | 21,192,701 | 453,698 | 695,758 | 242,061 (216,201 to 267,526) | 53 (48 to 59) | 1.14 |
| 1918 | Sweden | 5,813,850 | 78,570 | 104,591 | 26,022 (20,898 to 30,895) | 33 (27 to 39) | 0.45 |
| 1918 | Switzerland | 3,871,870 | 50,256 | 75,034 | 24,779 (21,958 to 27,548) | 49 (44 to 55) | 0.64 |
| 1957 | Spain | 29,499,414 | 280,210 | 293,502 | 13,292 (-3,401 to 28,970) | 5 (-1 to 10) | 0.05 |
| 1957 | Sweden | 7,388,611 | 70,356 | 73,132 | 2,775 (179 to 5,316) | 4 (0 to 8) | 0.04 |
| 1957 | Switzerland | 5,130,100 | 52,027 | 51,066 | -961 (-3,812 to 1,632) | -2 (-7 to 3) | -0.02 |
| 2020 | Spain | 47,353,706 | 422,004 | 492,930 | 70,926 (50,451 to 90,911) | 17 (12 to 22) | 0.15 |
| 2020 | Sweden | 10,379,295 | 90,199 | 97,870 | 7,671 (4,247 to 11,013) | 9 (5 to 12) | 0.07 |
| 2020 | Switzerland | 8,636,560 | 66,093 | 75,570 | 9,478 (6,380 to 12,482) | 14 (10 to 19) | 0.11 |

Expected=Expected all-cause deaths
Observed=Observed all-cause deaths
Excess=Excess all-cause deaths
% Excess=Percentage excess all-cause deaths as compared to expected all-cause deaths
CrI=credibility interval.
% Excess Pop=Excess all-cause deaths in per cent of the population

**Figure 1:**
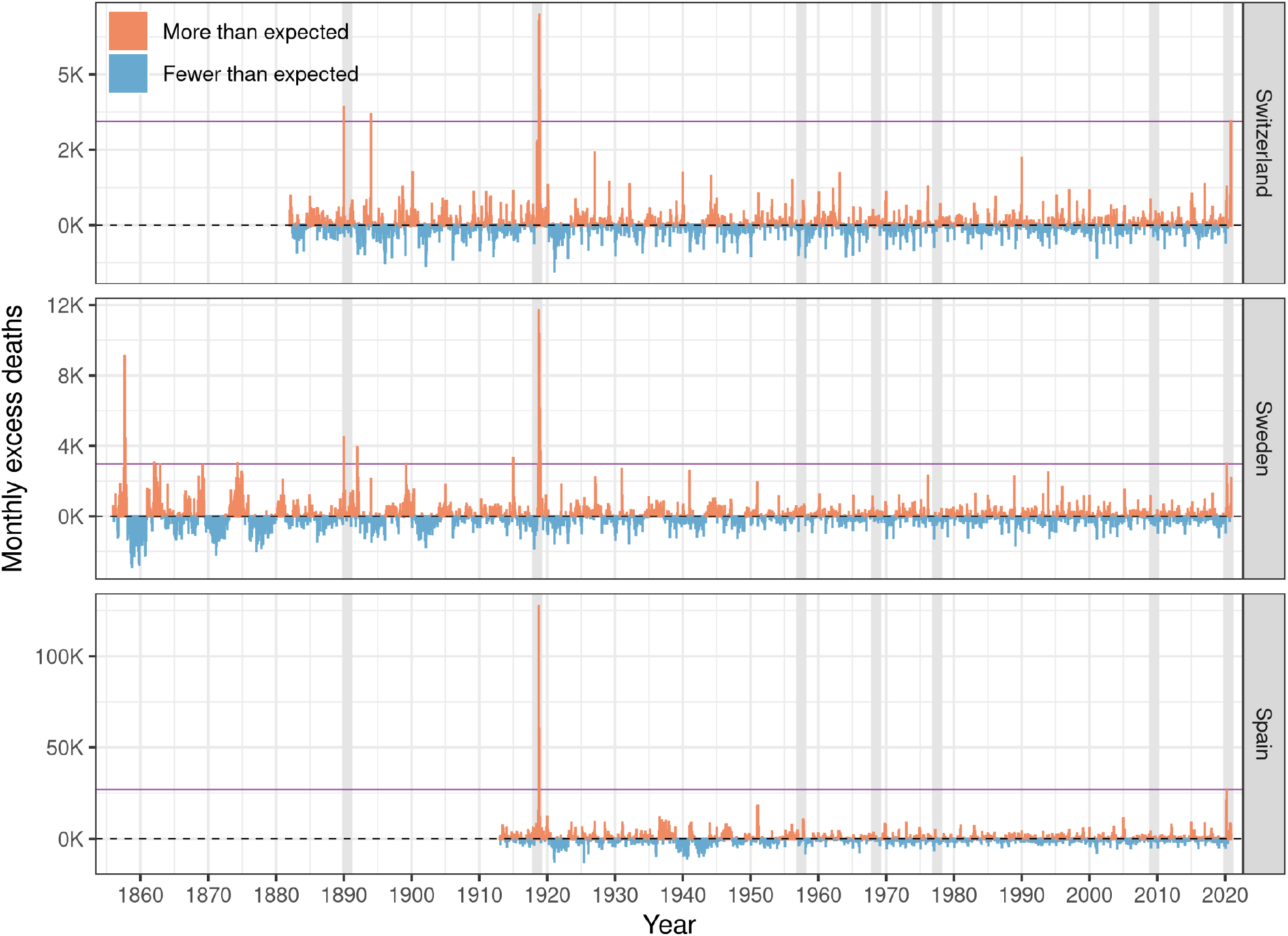
Monthly numbers of all-cause excess deaths in Spain, Sweden, and Switzerland displayed as differences between observed and expected values. The purple horizontal line marks the level of the highest monthly peak in 2020 for each country. Pandemic years are shaded in grey. The y-axes for the 3 countries have a different scaling to make them comparable in size.

Our detailed inspection of the 4 deadliest pandemic years (1890, 1918, 1957, and 2020) from the last 140 years revealed that the 1890 influenza led to marked excess mortality in Sweden and Switzerland during the month of January only (Fig. 2). The 1957 influenza pandemic led to only moderate excess mortality for the last 2-4 months of the year in all 3 countries. In contrast, excess mortality was very high during the 1918 influenza pandemic in the autumn for all 3 countries, and particularly high for Spain. In Switzerland, there were 2 waves between July and December 1918, while in Sweden and Spain there were significant single waves in the last quarter of 1918. The COVID-19 pandemic in 2020 consisted of 2 waves that affected the 3 countries differently, depending on the time of year. Although excess mortality was higher during the spring than for the autumn wave in Spain, it was the opposite in Switzerland. Sweden recorded similar excess mortality in the spring and autumn of 2020.

**Figure 2:**
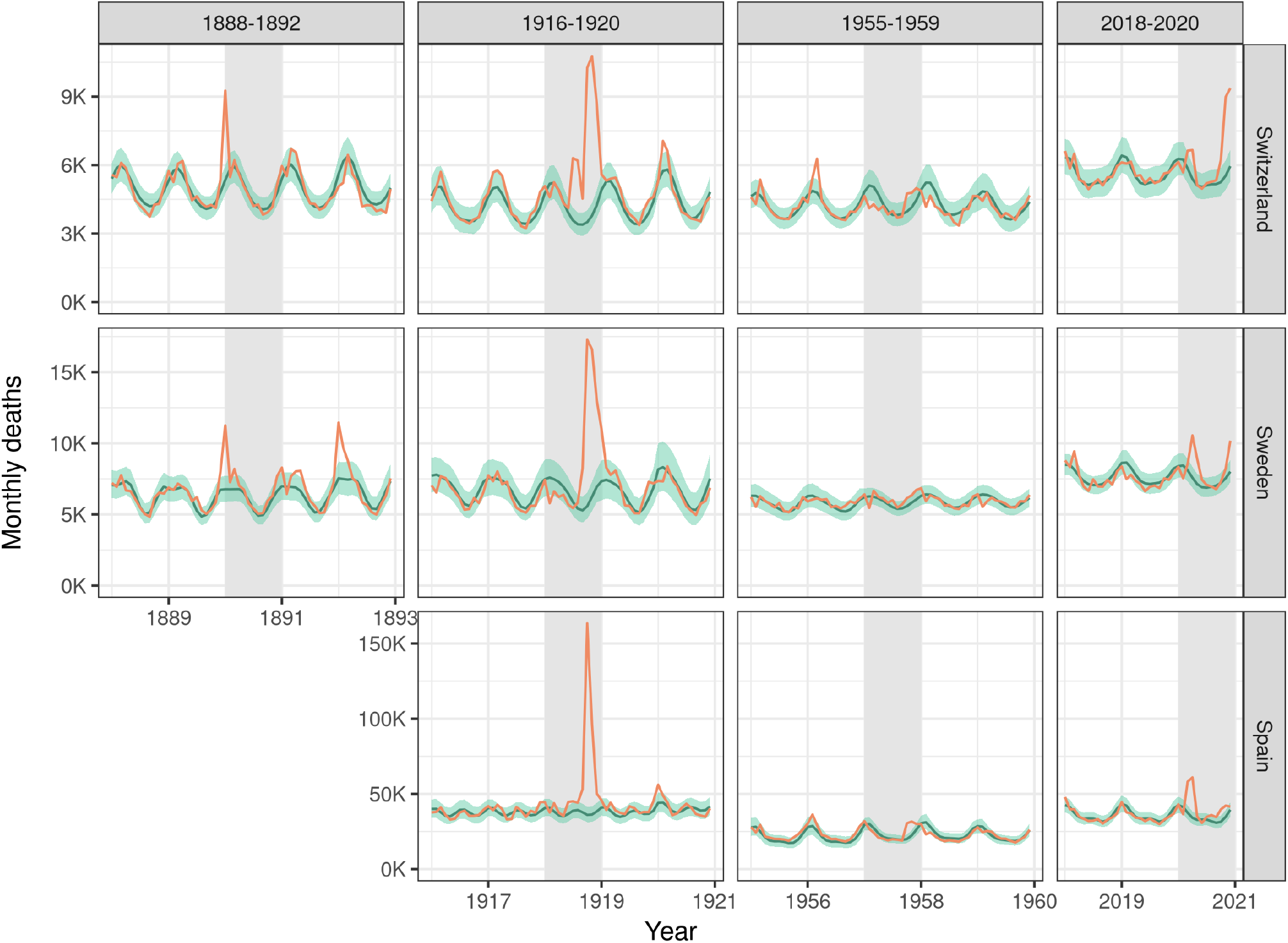
Detailed inspection of the strongest pandemic years 1890, 1918, 1957, and 2020 for all 3 countries. The black lines and grey areas indicate expected monthly all-cause deaths with 95% credible intervals based on the previous 5 years (excluding pandemic years themselves); the red lines are the observed monthly deaths. Pandemic years are shaded in grey. The y-axes for the 3 countries have a different scaling to make them comparable in size.

The annual excess deaths per age group (Fig. 3) revealed that the influenza pandemics of 1890 and 1957 affected all age groups relatively homogeneously. In contrast, we see mortality marked age patterns for 1918 and 2020. In 1918, young adults died much more frequently than in previous years (about 100 to 300% more deaths than expected for people aged 10-40 in all 3 countries). In contrast, in 2020, older adults (aged 70 and older) were most affected in all 3 countries. In both cases, the figures for the pandemic years for the respective age groups are clearly higher than for the previous 5 years in all 3 countries.

**Figure 3:**
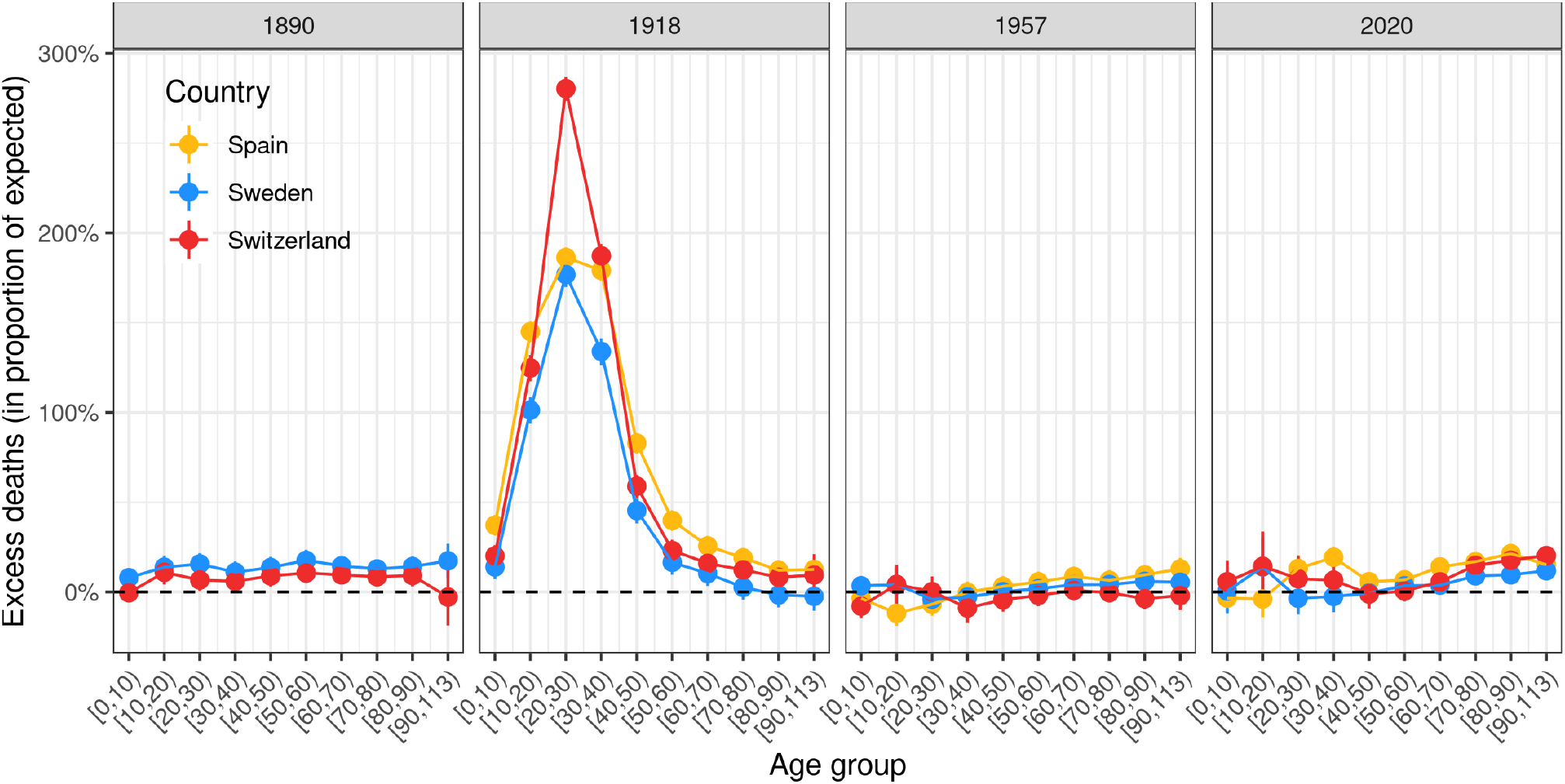
Excess all-cause deaths by age group for deadliest pandemic years 1890, 1918, 1957, and 2020 for all 3 countries. The number of excess deaths with 95% credible intervals are scaled by the number of expected deaths in each age group.

The concept of excess mortality has frequently been used to assess the overall impact of historical pandemics on specific populations. For example, international comparisons have been made for specific pandemics (*22*–*26*), and different historical pandemics have been compared with each other for particular countries (*27, 28*). Recently, a few studies based on non-continuous data have been published comparing excess mortality in 2020 with 1918 (*29, 30*). A study comparable to ours based on weekly data focused exclusively on Sweden and showed similar results (*31*).

On the one hand, our analysis of excess mortality in these 3 countries during 2020 confirms previous studies that have either made country comparisons (*3, 14, 32, 33*) or separately analyzed data for these countries (*31, 34*–*36*). Yet, on the other hand, our study contributes to scholarship in historical demography that examines longitudinal changes in mortality. However, most of these prior studies focus on annual data and do not include recent years, including 2020 (*37, 38*). To our knowledge, our study is the first to analyze a continuous series of monthly death data spanning more than 100 years, including 2020, that incorporates age composition and population structures.

Our work also points to a topical methodological question: Should extreme pandemic years be included in calculations of expected values for subsequent years or not (as we did here)? This is related to the question of whether one wants to give a clear counterfactual meaning to the term “expected”. If expected is meant to describe what would be observed without a pandemic, then evidently, expected mortality for a pandemic’s second year should not factor in deaths from the first year. For instance, in the years following 1918 we show that this decision makes a difference (Supplementary Fig. S3). This decision becomes more complicated for longer-termed comparisons when considering countries where deaths rose sharply over several years during world wars (*39*).

During the 2009 pandemic and especially during the ongoing 2020 pandemic, public and scientific interest in prior pandemics has increased markedly (*40*). However, comparisons with prior pandemics must be made with caution since there are parallels but also important differences (*41*). Lessons from the past cannot be transferred onto current challenges without nuanced understandings. When comparing the 2020 with the 1918 pandemic, it must be emphasized that the historical living and material context during and after the First World War differed substantially from now. Also, the virus responsible for each pandemic had different biological and epidemiological characteristics, including mortality patterns (*41*). Yet, there are also significant parallels between the 2 pandemics. For instance, authorities’ management of subsequent waves as recently described for Switzerland (*42*).

The 3 countries we studied have largely been spared high mortality effects from pandemics between 1918 and 2020. We postulated a pandemic disaster memory gap inspired by natural disaster research (*43, 44*) within this context. For certain populations within geographic locations, the absence of recent epidemics might have caused a loss of pandemic disaster memory; and, consequently, an increasing disregard of immediate epidemic risks in the wider population and among policymakers. That means that experiences from prior epidemics are insufficiently present in the public at large. To a certain extent, this is also reflected in pandemic preparedness. For example, the Swiss Influenza Pandemic Plan insufficiently addressed subsequent waves and their specific challenges (*45*).

Our study has important limitations. First, national sex-, age-, and cause-specific monthly death statistics were not available for certain years during the 20^th^ century (*46*) but our methodology partially accounted for this. Second, current analyses from the COVID-19 pandemic show that not all regions within these 3 countries were equally affected (*36, 47*). For this reason, spatial aggregation at the country level may hide regional nuances that are possibly important; however, historical data for such comparisons are scarce. Third, although our observed values for deaths in 2020 are in good agreement with officially reported figures from all 3 countries, there may be minor discrepancies in the expected values and, therefore, minor discrepancies with the absolute and relative excess mortality with other recent publications (*1, 14*). These dissimilarities might be due to methodological variations (e.g., full calendar year versus other time periods or only adding excess mortality and not mortality deficits) (*48*). Fourth, we did not aim to quantify the overall mortality impact from the civil war in Spain; instead, we quantified infectious disease mortality impact. The impact of the civil war in Spain is difficult to assess because the official death figures might be underestimated (*49*). However, suppose we apply the same methods as previous studies (*21*) with the official data used here. In that case, we arrive at a similar magnitude of several hundred thousand excess deaths from 1936-1939. Fifth, focusing on deaths does not account for other direct or indirect health, societal, or economic consequences during pandemic periods (*50*).

In sum, our results feature relevant historical dimensions of the COVID-19 pandemic. In Switzerland, historians have classified the 1918 influenza pandemic as the largest demographic disaster of the 20th century (*51*). Following the evidence on excess mortality, the 2020 pandemic led to the second largest demographic disaster in over 100 years in the three countries we studied. Looking beyond these 3 countries to other European ones shows that such excess mortality in 2020 was by no means inevitable. For example, Northern European countries such as Finland, Norway, and Denmark have not reported excess mortality (*14*). The exact reasons for these differences have yet to be investigated. However, the type, duration, and strength of governmental, public health, and direct infection control interventions, as well as ecological and cultural factors, might have played roles worthy of further study.

## Data Availability

All data and code used in this analysis are provided here: https://github.com/RPanczak/ISPM_excess-mortality/.

## Author statements

## Acknowledgements

The authors thank Alfredo Morabia, Graham Mooney, Thomas E. Ewing, Frank Rühli, Oliver Grübner, Olivia Keiser, Miguel Hernan, Kiko Llaneras, and Kristin Marie Bivens for helpful comments and support. Calculations were performed on UBELIX (http://www.id.unibe.ch/hpc), the HPC cluster at the University of Bern.

## Funding

Foundation for Research in Science and the Humanities at the University of Zurich (grant STWF-21-011, K.S. and J.F.)

Swiss National Science Foundation (SNSF project number 189498, M.E.).

## Author contributions

Conceptualization: K.S., R.P., J.R., and M.Z.

Data curation: K.S., R.P., K.M., C.B., C.J., R.W., and J.F.

Formal analysis: R.P., M.Z., J.R., and K.M.

Funding acquisition: K.S. and M.E.

Investigation: K.S., R.P., K.M., S.E.M., J.F., J.R., and M.Z.

Methodology: K.S., R.P., J.R., and M.Z.

Resources: K.S., M.Z., and M.E.

Supervision: K.S., M.Z., M.E., and J.R.

Visualization: R.P., J.R., K.M., and M.Z.

Writing – original draft: K.S., R.P., J.R., and M.Z.

Writing – review & editing: All authors.

## Competing interests

The authors declare no competing interests.

## Data and materials availability

All data and code used in this analysis are provided in the online Supplementary Material and https://github.com/RPanczak/ISPM_excess-mortality

## Notes

### Competing Interest Statement

The authors have declared no competing interest.

### Funding Statement

This work was funded by the Foundation for Research in Science and the Humanities at the University of Zurich (grant STWF-21-011, grantees K.S. and J.F.) and the Swiss National Science Foundation (SNSF project number 189498, grantee M.E.).

### Author Declarations

This study re-uses publicly available and monthly aggregated mortality figures at the national level, published by the national statistical authorities. Therefore, ethics approval was not necessary in this case.

