## Supplementary Materials for "Pandemic excess mortality in Spain, Sweden, and Switzerland during the COVID-19 pandemic in 2020 was at its highest since 1918"

August 12, 2021

**2 Supplementary figures** 4

### 1 Material and methods

#### 1.1 Data sources and availability

Monthly all-cause death numbers for Switzerland (1877-2020) and Sweden (1851-2020) were provided by the Swiss Federal Statistical Office* and Statistics Sweden†, they are freely available. For Spain, the monthly death figures 1941-2020 were accessible through the Spanish Statistical Office, the years 1908-1940 had to be transcribed from historical reports provided on their webpage ‡. It should be noted that all statistical authorities still consider the 2020 figures as provisional.

The annual population numbers as well as annual deaths by age group (in 1-year bands) were obtained from the Human Mortality Database (HMD, www.mortality.org). Most recent annual death counts (2019-2020 for Spain and Switzerland; 2020 for Sweden) and population structure (2020 for Spain) unavailable in HMD were obtained from provisional publicly accessible data repositories of the national statistics services of the three countries. In case of Switzerland we used 2019 population structure due to lack of reasonable estimates for 2020. All raw and prepared data as well as R code used in this analysis are provided in the study's GitHub repository §.

#### 1.2 Data preparation

Monthly time series for each country until 2018 (Switzerland and Spain) or 2019 (Sweden) were combined with the provisional estimates from each country in most recent years. Yearly deaths by 1-year age groups were combined into 10-year age bands.

#### 1.3 Statistical model

We built a model where the linear predictor of deaths count in year $i$, month $j$ and age group $k$ $D_{i,j,k}$ depended on (1) an intercept by age group $\alpha_k$, (2) a yearly linear trend $\beta_1$, (3) a periodic component $\beta_{2,\dots,5}$ based on four cosine and sine functions to account for temporal trends and seasonal variability in mortality, and (4) the total population in the age group $P_{i,k}$ included as an offset:

$$D_{i,j,k} = \alpha_k + \beta_1 i + \beta_2 \sin\left(\frac{2\pi j}{12}\right) + \beta_3 \sin\left(\frac{4\pi j}{12}\right) + \beta_4 \cos\left(\frac{2\pi j}{12}\right) + \beta_5 \cos\left(\frac{4\pi j}{12}\right) + \log(P_{i,k}) \quad (1)$$

The quantities $D_{i,j,k}$ were treated as latent variables, since deaths counts were not available with this level of precision. Rather, available data consisted of overall death counts by month $\mathbb{M}_{i,j}$ and age group-specific death counts by year $\mathbb{A}_{i,k}$. The model was jointly fitted to both types of data. The sum of $D_{i,j,k}$ by month was fitted to overall death counts by month with a negative binomial likelihood:

$$M_{i,j} = \sum_k D_{i,j,k} \quad (2)$$

$$\mathbb{M}_{i,j} \sim \text{neg-bin}\left(M_{i,j}, \phi\right) \quad (3)$$

where $\phi$ is the overdispersion parameter. Simultaneously, the sum of $D_{i,j,k}$ by age group was fitted to age group-specific death counts by year with a multinomial likelihood:

$$A_{i,k} = \sum_j D_{i,j,k} \quad (4)$$

$$N_i = \sum_j A_{i,k} \quad (5)$$

$$\mathbb{A}_{i,k} \sim \text{multinom}\left(N_i, \frac{A_{i,k}}{N_i}\right) \quad (6)$$

The joint likelihood can thus be expressed as:

$$\Pr(\mathbb{M}, \mathbb{A} | \alpha, \beta_1, \dots, \beta_5, \phi) = \prod_{i,j} \text{neg-bin}\left(\mathbb{M}_{i,j} | M_{i,j}, \phi\right) \times \prod_{i,k} \text{multinom}\left(\mathbb{A}_{i,k} \middle| N_i, \frac{A_{i,k}}{N_i}\right) \quad (7)$$

*Swiss Federal Statistical Office, Todesfälle nach Monat und Sterblichkeit seit 1803 (2021), (available at https://www.bfs.admin.ch/bfs/de/home/statistiken/kataloge-datenbanken/daten.assetdetail.14387168.html).

†Statistics Sweden (SCB), Births and deaths per month by sex. Year 1851-2020 (2021), (available at https://www.statistikdatabasen.scb.se/pxweb/en/ssd/START__BE__BE0101__BE0101G/ManadFoddDod/).

‡Instituto Nacional de Estadistica (INE), Anuarios Estadísticos (2021), (available at https://www.ine.es/inebaseweb/libros.do?tntp=25687).

§https://GitHub.com/RPanczak/ISPM_excess-mortality/

This approach of "stratification and joint likelihood" was inspired by a recent work focusing on the COVID-19 infection-fatality ratio [1].

The model was implemented in Stan (the full code is available in the textttstan folder in study's GitHub repository) [2]. We selected weakly informative prior distributions for all model parameters [3, 4], that is normal distributions with mean 0 and standard deviation 10 for intercept and yearly and seasonal slope parameters and a Cauchy distribution with location 0 and scale 5 for the inverse of the overdispersion parameter. We estimated the posterior distributions with Stan's default Hamiltonian Monte Carlo algorithm [5] by sampling 1000 iterations after 1000 iterations for warm-up in four independent chains. We assessed convergence using the Gelman-Rubin convergence diagnostic and sampling efficiency using effective sample size. These diagnostics did not reveal any issues with mixing and convergence in all settings.

#### 1.4 Estimating excess deaths

The procedure to obtain excess deaths for year $i$ (by month or by age group) was as follows:

1. We fitted the model to the five previous years $\{i-5,\ldots,i-1\}$ and obtained posterior samples for all parameters.

2. From these posterior samples, we computed expected values of $D_{i,j,k}$ for year $i$ using equation 1.

3. We then summed these quantities by month or age group (equations 2 or 4), and drew values from the corresponding probability distributions (equations 3 or 6), obtaining a set of expected values of death counts for year $i$ by month or by age group.

4. We then subtracted the observed number of deaths on year $i$, and summarized the samples by their mean and 2.5% and 97.5% quantiles, obtaining point estimates and 95% credible intervals of excess deaths for year $i$ by month or by age group.

This Bayesian approach ensured a full propagation of uncertainty from the data into the expected values and thus the estimates of excess deaths. Since five years of data is needed to calculate expected counts the estimates of excess deaths are available five years after the start of data collection in each country. We first calculated expected deaths excluding data from pandemic years of 1890, 1918 and 1957, and then also calculated expected deaths using all available data (shown by the example of 1918 in Supplementary Figure S3).

#### 1.5 Monthly vs. weekly data

A comparison with the same seasonality adjustments between weekly[¶] and monthly aggregated calculation of excess mortality (unadjusted for age) for Switzerland, Spain, and Sweden showed small differences in the expected number of deaths (between -0.4% and +0.9%) for the full calendar years where the data was available between 2005 and 2020 (Supplementary Figure S4).

[¶]Short-term Mortality Fluctuations (STMF) data series, (available at `https://www.mortality.org/`).

#### 2 Supplementary figures

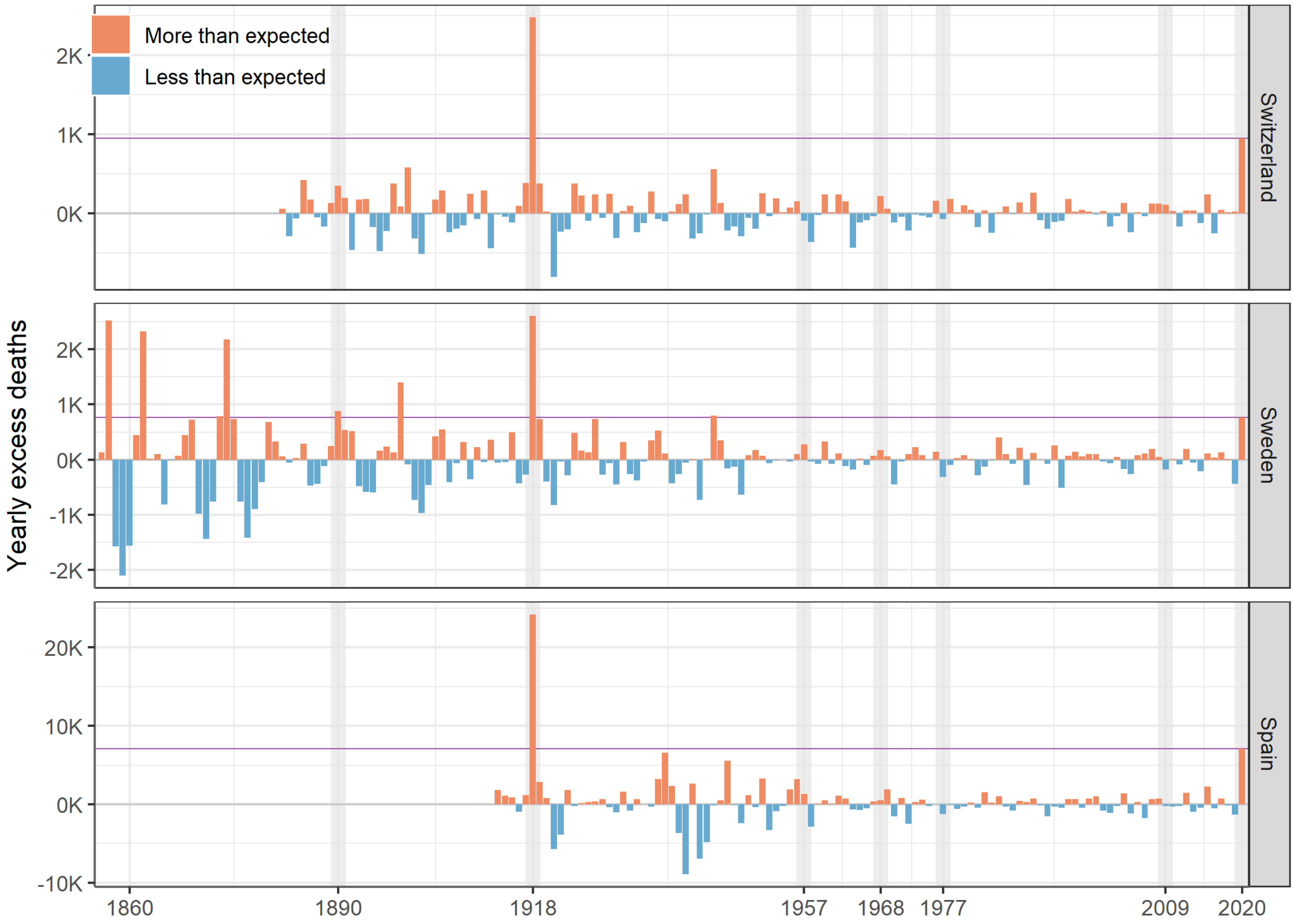

Figure S1: Annual numbers of deaths in Spain, Sweden, and Switzerland, displayed as differences between observed and expected number of deaths (red=more, blue=less). The purple horizontal line marks the level of 2020.

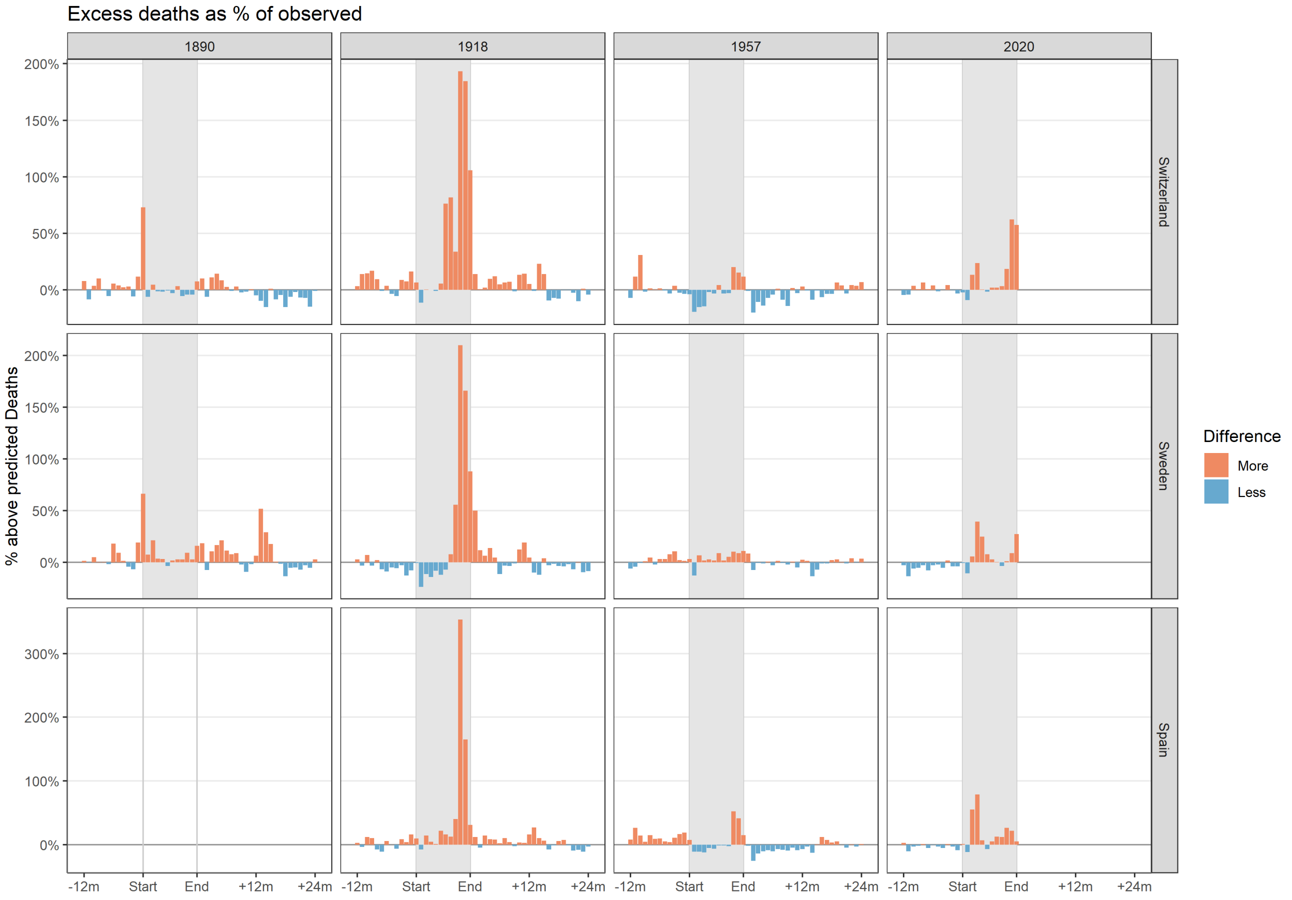

Figure S2: Detailed inspection of the strongest pandemic years 1890, 1918, 1957, and 2020 in all three countries. The differences between observed and expected values (red=more, blue=less) are displayed as percentages.

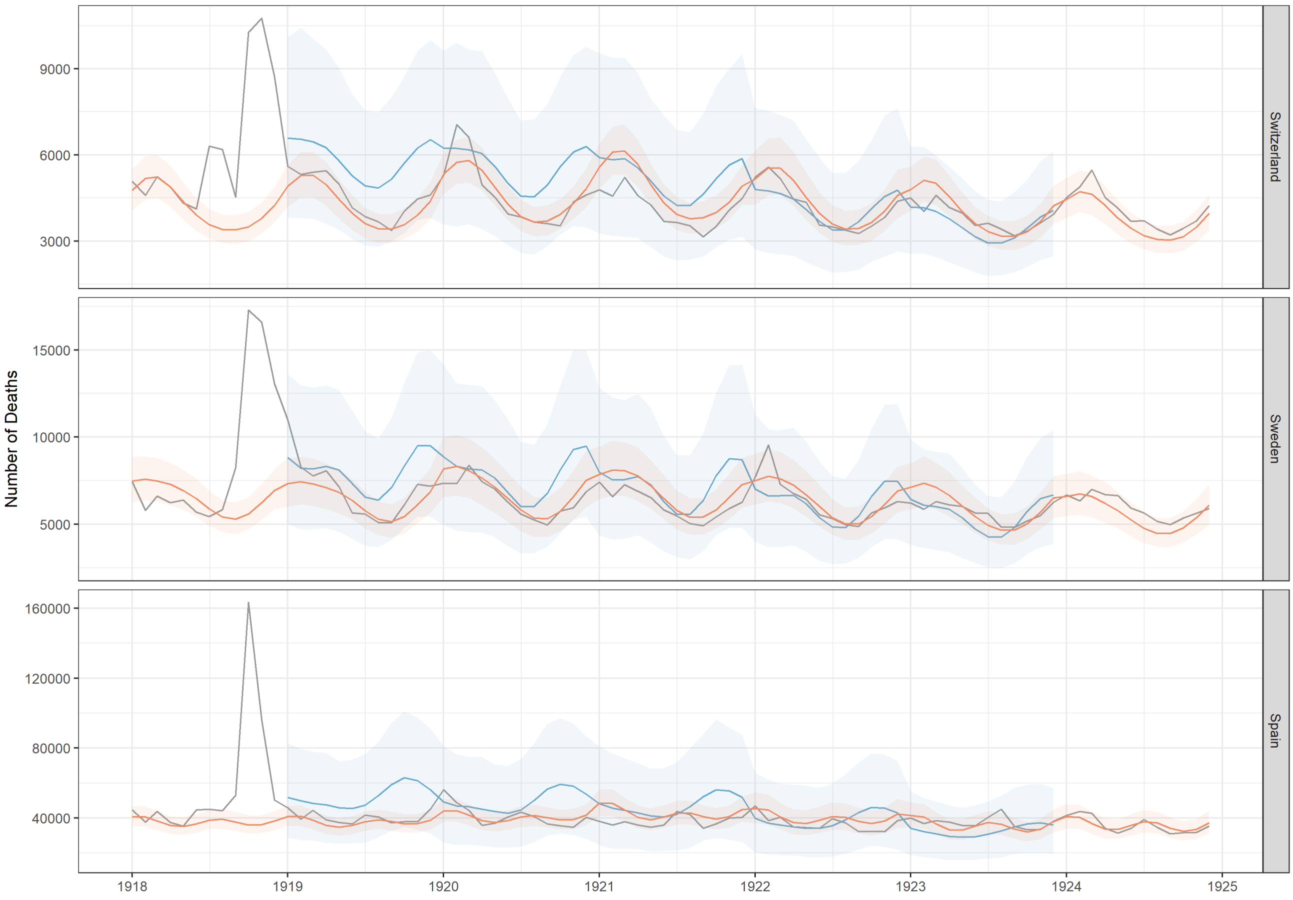

Figure S3: Exemplary visualization based on 1918, how big the difference in the calculated expected number of deaths is for the years 1919 and following, if the year 1918 is included in the calculations (blue line and confidence interval) or not (red line and confidence interval) (Black line: observed deaths). From 1924 onward the results of two methods are the same.

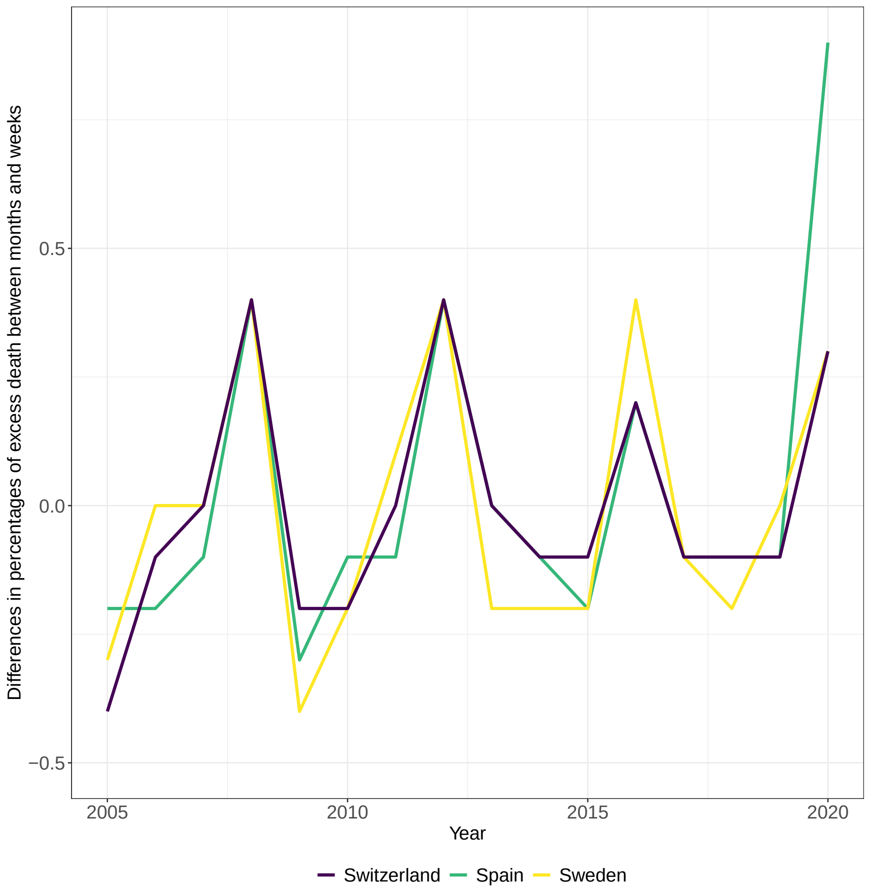

Figure S4: Comparison of weekly (STMF data from Human Mortality Database) vs. monthly (data as described and used in the main analyses) aggregated calculation of excess mortality for Switzerland, Spain, and Sweden and entire calendar years between 2005 and 2020. The differences between weekly and monthly calculated percentages of excess mortality range from -0.4% to 0.9%.
